# Distinct Longitudinal Clinical-Neuroanatomical Trajectories in Parkinson’s Disease Clinical Subtypes: Insight Towards Precision Medicine

**DOI:** 10.1101/2024.09.23.24314220

**Authors:** Seyed-Mohammad Fereshtehnejad, Roqaie Moqadam, Houman Azizi, Ronald B. Postuma, Mahsa Dadar, Anthony E. Lang, Connie Marras, Yashar Zeighami

**Affiliations:** Division of Neurology, Toronto Western Hospital, University of Toronto, Toronto, ON, Canada; The Edmond J. Safra Program in Parkinson’s Disease and the Morton and Gloria Shulman Movement Disorders Clinic, Toronto Western Hospital, UHN, Toronto, ON, Canada; Department of Neurobiology, Care Sciences and Society (NVS), Karolinska Institute, Stockholm, Sweden; Douglas Mental Health University Institute, Department of Psychiatry, McGill University, Montreal, QC, Canada; Integrated Program in Neuroscience, McGill University, Montreal, QC, Canada; Department of Neurology and Neurosurgery, Montreal Neurological Institute, McGill University, Montreal, QC, Canada

## Abstract

**Background:** Parkinson’s disease (PD) varies widely across individuals in clinical manifestations and course of progression. Identification and characterization of distinct biological subtypes could help explain this heterogeneity, identify the underlying pathophysiology, and predict disease progression across the subgroups of PD.

**Objective:** We aimed to compare long-term trajectories of various motor and non-motor clinical features, as well as patterns of brain atrophy between PD subtypes, using longitudinally acquired brain MRIs.

**Methods:** Data on 421 individuals with early-stage PD was retrieved from the Parkinson’s Progression Markers Initiative (PPMI), with an average follow-up time of 8.2 years until February 2024. Participants were classified into three clinical subtypes at the *de novo* stage using a previously validated subtyping criteria based on major motor and non-motor classifiers (early cognitive impairment, REM sleep behavior disorder (RBD), dysautonomia): ‘mild-motor predominant’ (n=223), ‘intermediate’ (n=146), and diffuse-malignant (n=52). To investigate the pattern of brain atrophy, we used T1-weighted MRIs from a subset of the PPMI population with at least two MRIs obtained, which consisted of 134 PD individuals and 60 healthy controls. Deformation-based morphometry (DBM) maps were calculated and mixed effect models were used to examine the interaction between PD subtypes and rate of atrophy across brain regions, controlling for sex and age at baseline.

**Results:** Compared to the ‘mild motor-predominant’ subtype, participants who were categorized as diffuse-malignant PD at baseline experienced greater worsening in motor severity (*p*=0.007), cognition (*p*<0.0001) and activities of daily living (ADL) (*p*<0.0001) after 8 years. Individuals with diffuse-malignant PD showed a significantly higher rate of atrophy across multiple brain regions, including precuneus, paracentral lobule, inferior temporal gyrus, fusiform gyrus, and lateral hemisphere of the cerebellum (corrected *p*<0.05).

**Conclusion:** Our study revealed a distinct pattern of long-term progression in various motor and non-motor clinical outcomes between different subtypes of idiopathic PD. Furthermore, we demonstrated an accelerated atrophy pattern within several brain regions in the diffuse-malignant PD subtype. These findings suggest a more widespread and aggressive neurodegenerative process in a subgroup of people with PD, favoring the existence of diverse underlying pathophysiology with clinical relevance for future precision medicine in PD.

## INTRODUCTION

Parkinson’s disease (PD) varies widely from person to person in terms of clinical manifestations and progression. Although the clinical and biological signature of PD may be unique to the individual, identification of commonalities characterizing disease subtypes can be one step forward towards precision medicine.^1^ Biomarker characterization of distinct subtypes can parse this heterogeneity, shed light on the underlying pathophysiology, and predict disease progression across the PD spectrum. We previously introduced and validated a data-driven approach to define three subtypes of idiopathic PD using multi-domain clinical criteria.^2,3^ According to this classification system, early development of three key nonmotor features - mild cognitive impairment, orthostatic hypotension, and REM sleep behavior disorder (RBD) - at the drug-naïve stage defines a distinct subtype of idiopathic PD called the ‘diffuse-malignant’ subtype with the most rapid disease progression.^3^ Several independent studies also demonstrated faster disease progression, earlier development of dementia and shorter survival in the diffuse-malignant subtype.^4,5^ Nevertheless, there is a dearth of knowledge on the long-term clinical trajectories, as well as underlying pathophysiological causes of faster disease progression in a subgroup of people with PD. Using cross-sectional data from diffusion-weighted imaging, more prominent disruption in various cortico-subcortical connectivity networks was found in the diffuse-malignant subtype.^6^ Another cross-sectional study demonstrated further disease-related brain atrophy at baseline in the diffuse-malignant subtype using deformation-based morphometry (DBM) of T1-weighted magnetic resonance imaging (MRIs).^7^ A longitudinal clinical and brain imaging study can monitor clinical trajectories and pathological changes in brain structure over time, offering insights into the dynamics of atrophy for each PD subtype that cross-sectional imaging cannot capture. Thus, the aim of this study was to: (i) compare long-term clinical trajectories of the main motor and non-motor features between PD subtypes defined at the early untreated stage, (ii) use longitudinally acquired brain MRIs to investigate and compare different patterns of brain atrophy between the PD subtypes (iii) and to compare the within and between subtype neuroanatomical features to establish convergence and divergence across their trajectories.

## METHODS

### Study Population

The Parkinson’s Progression Markers Initiative (PPMI) is a multicenter longitudinal cohort of individuals with early idiopathic Parkinson’s disease, untreated at recruitment^8^ (Detailed information at *http://www.ppmi-info.org*). The inclusion criteria are: age at least 30 years old, diagnosed with Parkinson’s disease within the past 2 years, at least two signs or symptoms of Parkinsonism (resting tremor, bradykinesia, and/or rigidity), a baseline Hoehn and Yahr Stage of I or II, and does not require symptomatic treatment within 6 months of baseline. For the purpose of this study, we obtained the T1 weighted MRIs from participants in whom at least two MRIs were obtained.

### Clinical Measures

The PPMI study conducted a thorough evaluation of motor and non-motor manifestations at the screening, baseline, and follow-up visit. For this study, we analyzed trajectories of the following clinical motor and non-motor disease domains over time:

i. Motor: sum of the severity scores for motor signs on the MDS-Unified Parkinson’s Disease Rating Scale (MDS-UPDRS) part III.^9^
ii. Cognition: global cognitive status measured by the Montreal cognitive assessment (MoCA) score.
iii. Autonomic dysfunction: total score of the Scales for Outcomes in PD-Autonomic (SCOPA-AUT) measuring gastrointestinal, urinary, cardiovascular, thermoregulatory, pupillomotor, and sexual functions.^10^ The SCOPA-AUT score ranges from 0 to 45, with higher scores indicating more severe autonomic dysfunction in PD.^10^
iv. Daily activities and functioning: total score of the Schwab and England activities of daily living (SE-ADL) score which measures functional independence in PD patients on a scale from 0 (complete dependence) to 100 (complete independence).^11^

### Subtyping Rules

Participants were classified into three clinical subtypes at the baseline early untreated stage using our previously validated multidomain subtyping criteria based on major motor (MDS-UPDRS-II and III) and non-motor classifiers (early cognitive impairment, REM sleep behavior disorder (RBD), dysautonomia).^3^ The following definitions were used to assign individuals to their clinical subtype: subtype I (mild motor-predominant): both composite motor score and all non-motor summary scores (NMS) below the 75^th^ percentile; subtype III (diffuse malignant): (composite motor score AND either >1 of three non-motor scores >75^th^ percentile, or all three non-motor scores >75^th^ percentile); and subtype II (intermediate): those individuals who do not meet criteria for subtype I or II.^2,3^

### Brain Imaging

We used T1-weighted magnetic resonance imaging (MRI) to evaluate the atrophy pattern in PD and control subjects over time. More specifically, we used deformation-based morphometry (DBM), which measures the local brain atrophy patterns using nonlinear transformation of the T1 weighted images to MNI-ICBM-2009c symmetrical template.^12^ These brain atrophy maps were averaged across 282 cortical and subcortical regions over time using the neuroanatomically defined regions by the Allen Institute for brain sciences’ atlas (*https://community.brain-map.org/t/allen-human-reference-atlas-3d-2020-new/405*) which provided us with the regional atrophy measures used in our statistical analysis.

### Neuroanatomical Heterogeneity

To examine the degree of within group and between group neuroanatomical heterogeneity, we used the regional DBM values explained in the previous section (i.e., 282 regions). We used the Spearman correlation between regional DBM values as a measure for similarity between two MRIs across all available subjects and visits with the baseline visits of participants (including controls) as the reference. For within-group similarity, we calculated the mean Fisher transformed correlation values between each visit of a given subject and all subjects within the same subtype at baseline. Similarly, for between-group similarity and to test whether the pattern of brain atrophy in mild motor-predominant and intermediate subtypes would converge over time to that of the diffuse-malignant subtype at baseline (i.e. significant increase in similarity to diffuse-malignant baseline compared to similarity to other groups). To do so, we used the mean Fisher transformed correlation values between each visit of a given subject and all subjects across the diffuse-malignant subtype at baseline.

### Statistical Analysis

For comparison of the demographics and baseline clinical features between the subtypes, we used one-way ANOVA or Chi square test. Next, we applied mixed effect models to examine clinical trajectories between PD subtypes over 8 years, using the following model:

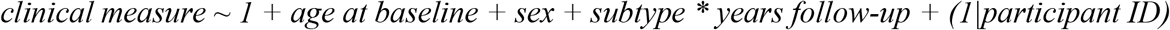

with subtype and its interaction with follow-up time as the main effect of interest, controlling for sex and age. The model evaluated longitudinal changes in MDS-UPDRS-III, MoCA, and SCOPA scores as our clinical measures of interest. Similarly, we used mixed effect models to examine (i) the atrophy pattern between each subtype and healthy control subjects and (ii) the interaction between PD subtypes and the rate of atrophy across brain regions over time, controlling for sex and age. Similarly, for neuroanatomical heterogeneity we used a mixed effect model using the following model:

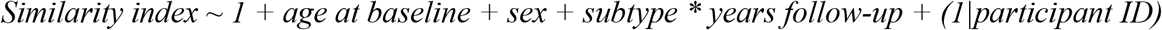

We used MATLAB R2023b for all statistical procedures and a 2-tailed p-value less than 0.05 was considered statistically significant. All results are reported after False Discovery Rate (FDR) correction for multiple comparison (FDR corrected p<0.05).

## RESULTS

### Demographics & Baseline Features

Data from 421 PD subjects (65.6% male) were used for clinical trajectory assessment. At baseline, 223 participants were categorized as mild motor-predominant, 146 as intermediate, and 52 as diffuse-malignant subtype. The average follow-up duration was 8.2±3.7 years, and 6481 total visits were included. **Table 1** summarizes demographics, motor and non-motor features within each PD subtype at baseline. As expected, and derived from the subtype definition, various motor and non-motor features were significantly more impaired at baseline in the diffuse-malignant subtype.

**TABLE 1.**
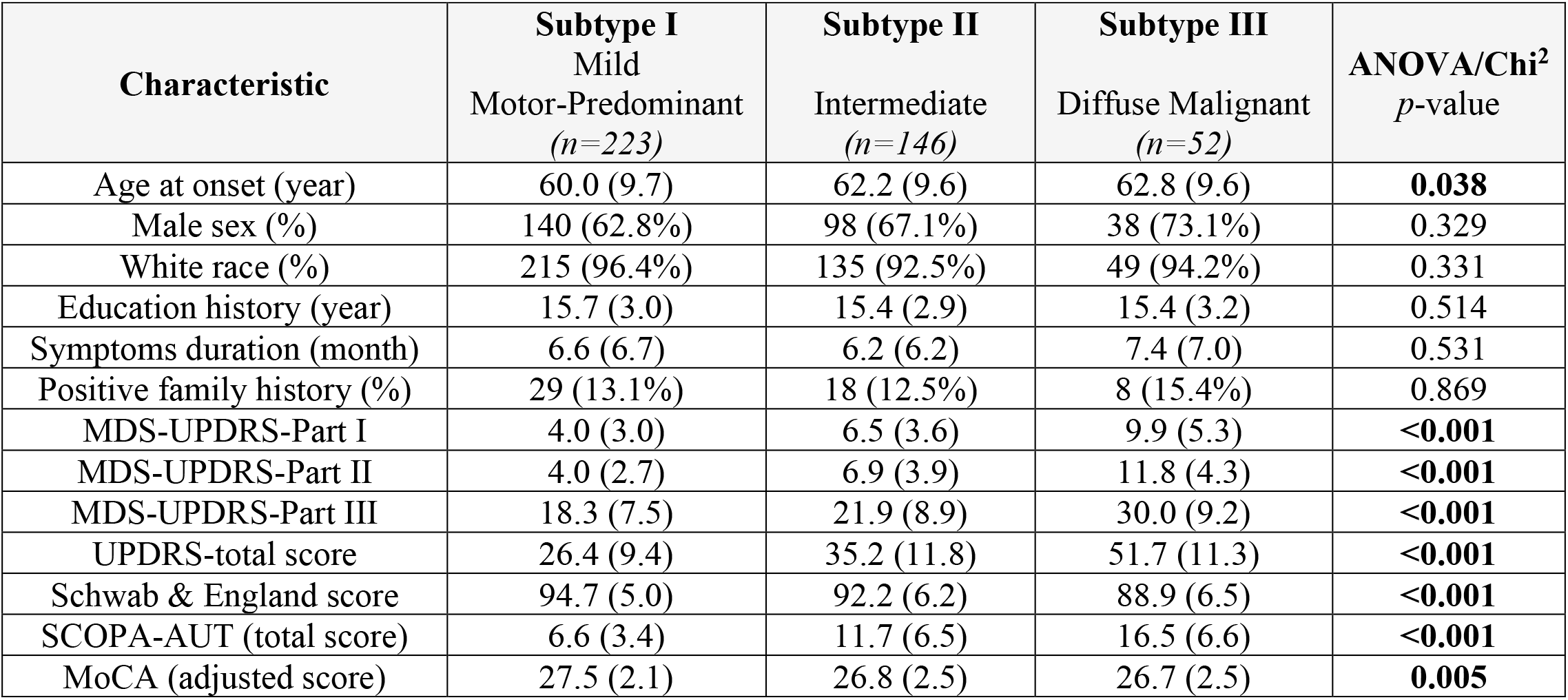
Demographics and baseline clinical features in individuals with different subtypes of Parkinson’s disease in the PPMI cohort (N=421)

| **Characteristic** | **Subtype I**<br>**Mild**<br>**Motor-Predominant**<br>**(n=223)** | **Subtype II**<br>**Intermediate**<br>**(n=146)** | **Subtype III**<br>**Diffuse Malignant**<br>**(n=52)** | **ANOVA/Chi<sup>2</sup>**<br>**p-value** |
| --- | --- | --- | --- | --- |
| Age at onset (year) | 60.0 (9.7) | 62.2 (9.6) | 62.8 (9.6) | **0.038** |
| Male sex (%) | 140 (62.8%) | 98 (67.1%) | 38 (73.1%) | 0.329 |
| White race (%) | 215 (96.4%) | 135 (92.5%) | 49 (94.2%) | 0.331 |
| Education history (year) | 15.7 (3.0) | 15.4 (2.9) | 15.4 (3.2) | 0.514 |
| Symptoms duration (month) | 6.6 (6.7) | 6.2 (6.2) | 7.4 (7.0) | 0.531 |
| Positive family history (%) | 29 (13.1%) | 18 (12.5%) | 8 (15.4%) | 0.869 |
| MDS-UPDRS-Part I | 4.0 (3.0) | 6.5 (3.6) | 9.9 (5.3) | **<0.001** |
| MDS-UPDRS-Part II | 4.0 (2.7) | 6.9 (3.9) | 11.8 (4.3) | **<0.001** |
| MDS-UPDRS-Part III | 18.3 (7.5) | 21.9 (8.9) | 30.0 (9.2) | **<0.001** |
| UPDRS-total score | 26.4 (9.4) | 35.2 (11.8) | 51.7 (11.3) | **<0.001** |
| Schwab & England score | 94.7 (5.0) | 92.2 (6.2) | 88.9 (6.5) | **<0.001** |
| SCOPA-AUT (total score) | 6.6 (3.4) | 11.7 (6.5) | 16.5 (6.6) | **<0.001** |
| MoCA (adjusted score) | 27.5 (2.1) | 26.8 (2.5) | 26.7 (2.5) | **0.005** |

### Clinical Trajectory

Mixed-effects models demonstrated important differences in the longitudinal trajectories of various outcomes (**Figure 1**). As listed in **Table 2**, compared to the mild motor-predominant subtype, individuals who were initially categorized as intermediate or diffuse-malignant subtypes at baseline experienced faster progression in motor severity, with 0.37 (95% CI: 0.21-0.53, *p*<0.0001) and 0.41 (95% CI: 0.11-0.71, *p*=0.0070) units additional annual increase in MDS-UPDRS-III score. A more rapid cognitive decline was also demonstrated in the diffuse-malignant subtype. After adjusting for baseline MoCA score and age, members of the diffuse-malignant subtype demonstrated -0.41 (95% CI: -0.51 to -0.32, p<0.0001) additional decline in MoCA score per year when compared to the mild motor-predominant subtype. A distinct trajectory was also noted in the overall impact of PD on independence in ADL. Individuals categorized as the intermediate or diffuse-malignant subtypes at baseline had a significantly faster decline in ADL compared to the mild motor-predominant subtype, with -0.59 (95% CI: -0.72 --0.47, p<0.0001) and -0.92 (95% CI: -1.14 --0.70, p<0.0001) percent further decline in SE-ADL score per year, respectively.

**TABLE 2.** Mixed-effects models to assess longitudinal trajectories of various clinical outcomes after >8 years of follow-up in individuals with different subtypes of Parkinson’s disease in the PPMI cohort (N=421)

|  | Effect Size | 95% CI | t Statistic | p-value |
| --- | --- | --- | --- | --- |
| **Outcome: Motor Severity (MDS-UPDRS-III)** |  |  |  |  |
| Age Effect | 0.20 | 0.12-0.29 | 4.85 | **0.0007** |
| Group*Time Interaction |  |  |  |  |
| Subtype-I: Mild Motor-Predominant | Reference | - | - | - |
| Subtype-II: Intermediate | 0.37 | 0.21-0.53 | 4.48 | **<0.0001** |
| Subtype-III: Diffuse Malignant | 0.41 | 0.11-0.71 | 2.70 | **0.0070** |
| **Outcome: Cognition (MoCA)** |  |  |  |  |
| Age Effect | -0.11 | -0.13 - -0.08 | -8.57 | **<0.0001** |
| Group*Time Interaction |  |  |  |  |
| Subtype-I: Mild Motor-Predominant | Reference | - | - | - |
| Subtype-II: Intermediate | -0.12 | -0.17 - -0.07 | -4.78 | **<0.0001** |
| Subtype-III: Diffuse Malignant | -0.41 | -0.51 - -0.32 | -8.65 | **<0.0001** |
| **Outcome: Autonomic Symptoms (SCOPA-AUT)** |  |  |  |  |
| Age Effect | 0.11 | 0.06-0.16 | 4.16 | **<0.0001** |
| Group*Time Interaction |  |  |  |  |
| Subtype-I: Mild Motor-Predominant | Reference | - | - | - |
| Subtype-II: Intermediate | -0.02 | -0.10 – 0.06 | -0.46 | 0.64 |
| Subtype-III: Diffuse Malignant | 0.14 | 0.00-0.29 | 1.91 | 0.055 |
| **Outcome: Daily Activities (Schwab & England-ADL)** |  |  |  |  |
| Age Effect | -0.12 | -0.18 - -0.06 | -3.78 | **0.0002** |
| Group*Time Interaction |  |  |  |  |
| Subtype-I: Mild Motor-Predominant | Reference | - | - | - |
| Subtype-II: Intermediate | -0.59 | -0.72 - -0.47 | -9.08 | **<0.0001** |
| Subtype-III: Diffuse Malignant | -0.92 | -1.14 - -0.70 | -8.12 | **<0.0001** |

CI: confidence interval; ADL: activities of daily living

**FIGURE 1.**
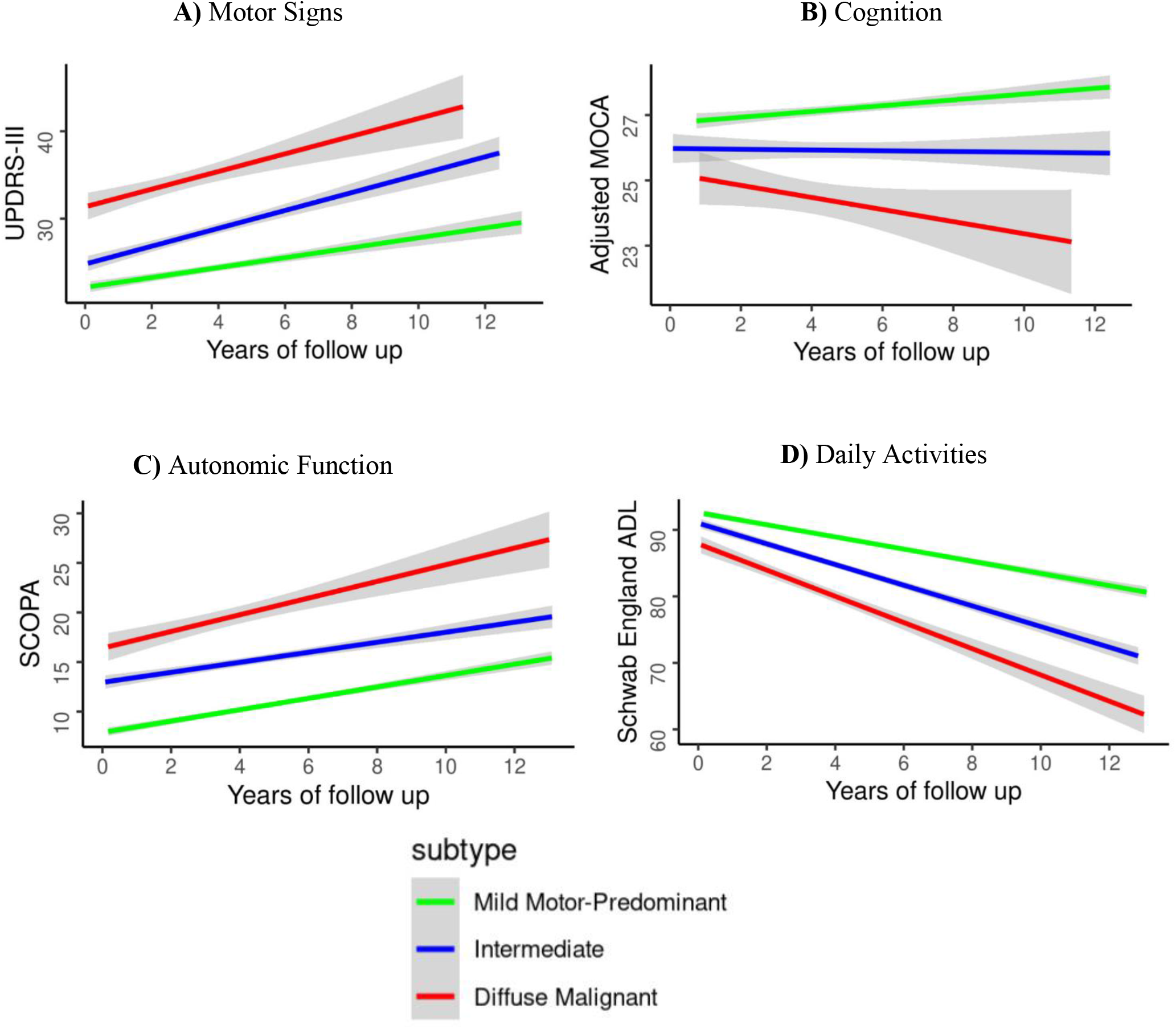
Distinct clinical trajectories in motor severity (A: UPDRS-III score), cognition (B: MOCA score), dysautonomia (C: SCOPA total score) and activities of daily living (D: Schwab and England ADL score) between clinical subtypes of PD after 8-10 years of follow-up (all mixed-effect models *p*-value > 0.001).

### Brain Atrophy Trajectory

For the neuroanatomical assessment, longitudinal MRI studies were available for a total of 194 participants, including 134 PD participants and 60 healthy controls comprising a total of 606 visits. Among the PPMI subpopulation included in the longitudinal MRI analysis, the sample size for each clinical subtype was as follows: mild motor-predominant (n=74), intermediate (n=44), and diffuse-malignant (n=16). The average time between the first and the last brain MRIs was 3.4±1.1 years in the PD cohort and 1.9±1.3 years in healthy controls.

All three groups showed significant atrophy over time in the midbrain including substantia nigra and red nucleus as well as the amygdala in comparison with healthy controls. In the direct comparison between subtypes, individuals with diffuse-malignant PD showed a significantly higher rate of atrophy in comparison to mild motor-predominant subtype across multiple brain regions [**Figure 2**, blue colored], including precuneus as well as inferior and middle temporal gyrus regions (i.e. hippocampal area) implicated in cognitive decline and dementias. We also found accelerated atrophy in the occipital regions of the cortex and cerebellar hemispheres in the diffuse-malignant subtype. Finally, we found accelerated enlargement of ventricles in the diffuse-malignant PD compared to mild motor-predominant PD in line with faster tissue loss. In contrast, we found bilateral further interval atrophy in the body of caudate in the mild motor-predominant compared to the diffuse-malignant group [**Figure 2**, red colored]. The intermediate subtype showed an accelerated atrophy only in the putamen region in comparison with the mild motor-predominant subtype after correction for multiple comparison.

**FIGURE 2.**
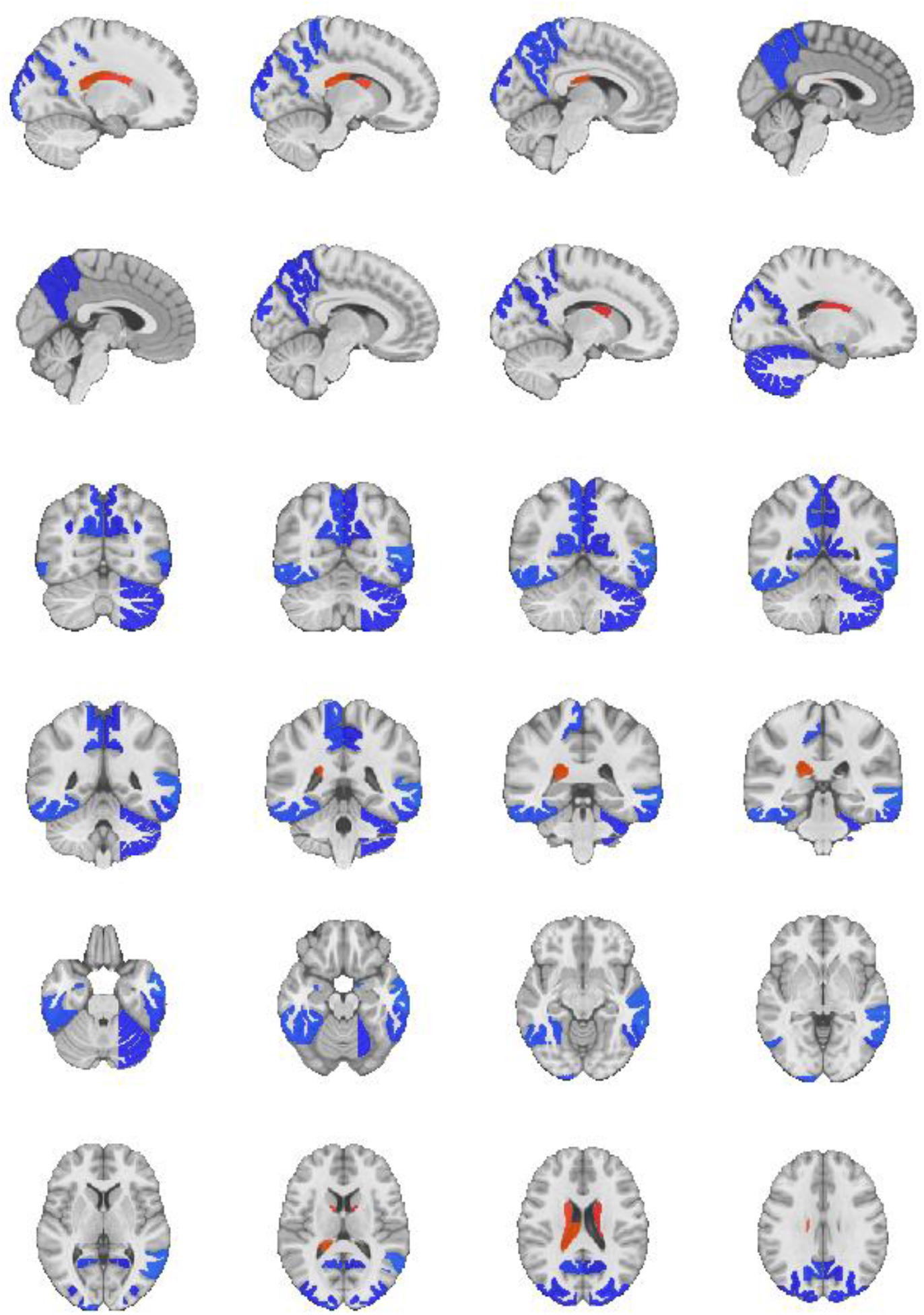
Significantly accelerated atrophy was observed in the diffuse-malignant subtype compared with the mild motor-predominant subtype. Blue colors show a higher negative slope in DBM signal in the diffuse-malignant subtype compared to the mild motor-predominant group, whereas warm colors show a less steep atrophy in the case of caudate, or a more positive slope in the case of ventricular enlargement. The results show a widespread acceleration in cortical regions including occipital cortex, inferior and middle temporal gyrus, as well as the precuneus. There is also faster atrophy observed in the cerebellar regions. There was a higher rate of atrophy in bilateral caudate in the mild motor-predominant subtype compared with the diffuse-malignant subtype. Finally, we observed an accelerated enlargement of ventricles in the diffuse-malignant subtype.

### Within And Between Subtypes Neuroanatomical Heterogeneity

As illustrated in **Figure 3-A**, based on the linear mixed effect model, the largest within subtype similarity in the pattern of brain atrophy was seen in the diffuse-malignant subtype with a statistically significant correlation between DBM values across brain regions (β=0.07, t=3.67, *p*=0.00026). To check whether there is an ultimate convergence in the pattern of brain atrophy between the subtypes (i.e. possibly suggesting similar final outcomes with the diffuse malignant group simply getting there more rapidly rather than distinct bio-anatomical subgroups), we evaluated between subtypes similarity by correlating longitudinal pattern of brain atrophy in the mild motor-predominant and intermediate subtypes with that of the diffuse-malignant subtype at baseline. As illustrated in **Figure 3-B**, after 3.4 years of follow-up, the increase in the correlation coefficient between the overall pattern of longitudinal brain atrophy in the mild motor-predominant and intermediate subtypes and the baseline pattern of brain atrophy in the diffuse-malignant subtype was not significantly different from the increase observed in the correlation coefficient over time between healthy controls and the baseline pattern of brain atrophy in diffuse malignant subtype (*p*=0.625 and *p*=0.411, respectively). However, subjects with diffuse-malignant subtype demonstrated an increase in overall correlation coefficient with their own baseline regional DBM values over time, which was significantly faster than that of the healthy controls (*p*=0.0203).

**FIGURE 3.**
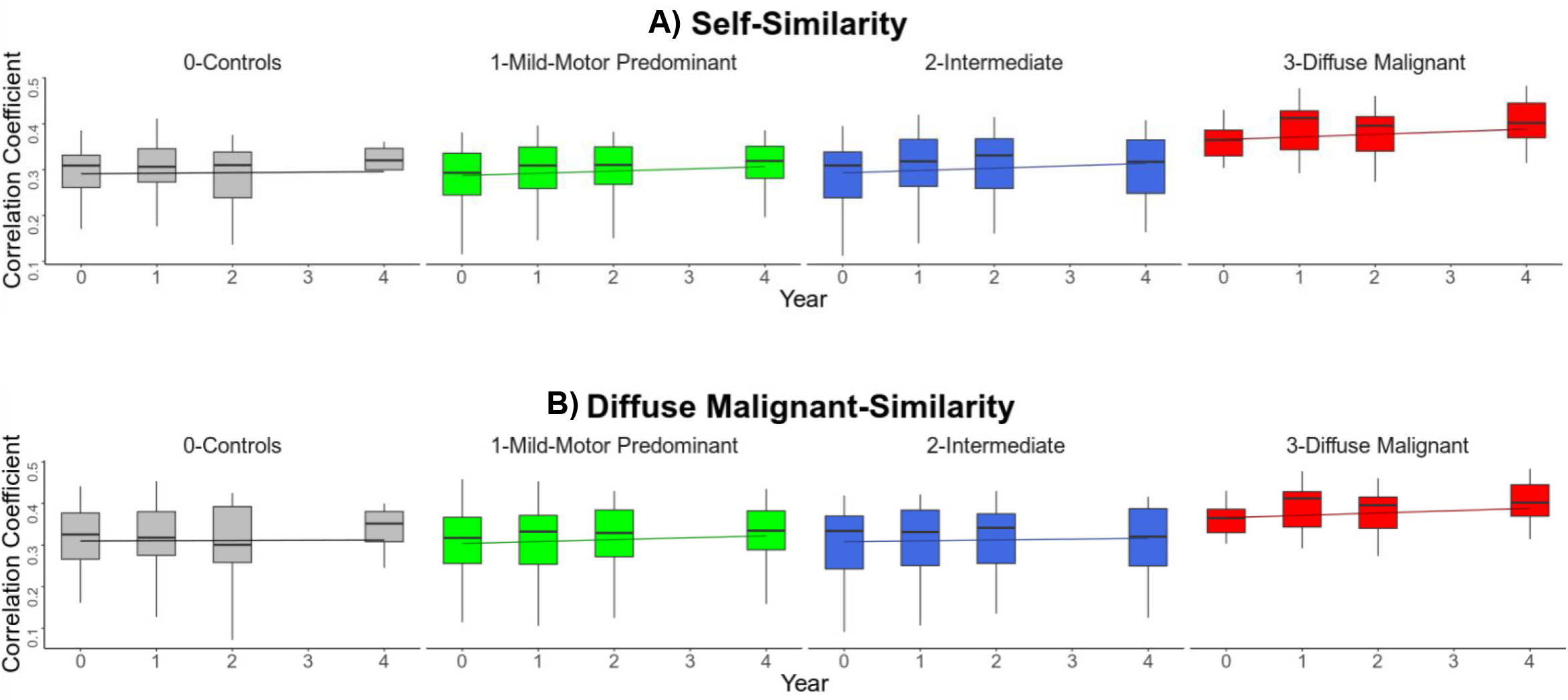
Spearman correlations and mixed effect models to assess within-group (self-similarity) and between-group (similarity to baseline diffuse-malignant pattern) neuroanatomical heterogeneity. In self-similarity analysis, based on the mixed effect model, members of diffuse-malignant subtype had the highest baseline within-group similarity in regional DBM values of brain atrophy (β=0.07, t=3.67, *p*=0.00026). Over 4-years of follow-up, the progression of brain atrophy in subjects with mild motor-predominant and intermediate subtypes though trending towards that of the diffuse-malignant subtype at baseline, was not significantly different compared to healthy controls (‘normal aging’) (β=0.000, t=-0.48, *p*=0.62, and β=-0.001, t=-0.82, *p*=0.411, respectively). On the other hand, members of the diffuse-malignant subtype showed a statistically significant increase in the correlation coefficient of diffuse malignant similarity over time when compared to the trend seen in healthy controls (*p*=0.0203).

## DISCUSSION

Our study revealed a distinct pattern of disease progression across subtypes of individuals with idiopathic PD. Participants initially subtyped as diffuse-malignant at the early stage, when dopaminergic treatment was not yet deemed necessary, had a significantly faster clinical progression in motor and non-motor domains during long-term follow-up. We discovered a hierarchical trend in the slope of progression in clinical measures between the subtypes after >8 years of follow-up. Specifically, individuals classified under the diffuse-malignant subtype displayed a faster decline in cognitive function and a more rapid progression of motor severity compared to those in the mild motor-predominant subtype. Consequently, these participants exhibited a faster decline in activities of daily living. Similarly, participants in the intermediate subtype demonstrated rates of decline falling between those of the ‘mild motor-predominant’ and diffuse-malignant subtypes. Even though differences in clinical measures exist at baseline, which are inherent to the multi-domain definition of the subtypes, these differences are augmented in longitudinal trajectories over follow-up with distinctive slopes of progression that remained statistically significant after adjustment for the effect of aging and baseline clinical differences. In fact, the most eccentric variation in longitudinal trajectory was observed in Schwab and England ADL, a global measure that is not used to define subtyping membership at the early untreated stage. Our findings are consistent with those of a few other longitudinal studies reporting a more aggressive disease trajectory in individuals with PD with severe motor and non-motor phenotypes at baseline and an older age of onset.^13,14^

Using longitudinal brain MRI scans, we demonstrated an accelerated atrophy pattern within several cortical regions and cerebellum in participants who were initially classified as having the diffuse-malignant PD subtype at the *drug-naïve* stage. This faster rate of brain atrophy in the diffuse-malignant subtype was evident even after a short follow-up period (approximately 3.5 years). In line with our findings, another study showed a more pronounced grey matter thinning in parahippocampal and transverse temporal gyri in individuals with early-stage PD who concurrently had severe olfactory dysfunction and RBD.^15^ Interestingly, some of the MRI regions with more atrophy in the diffuse-malignant subtype of idiopathic PD overlap remarkably with those of regions with cholinergic terminal loss in dementia with Lewy bodies (DLB) as shown by PET imaging.^16^ These findings suggest the presence of a more diffuse multidomain neurodegenerative process in a subgroup of people with PD, the diffuse-malignant subtype, with more expanded underlying pathophysiology, probably with more prominent multi-proteinopathies and earlier multi-neurotransmitter system involvement. A postmortem study demonstrated evidence for different rates of progression of Lewy pathology and Alzheimer’s-related pathology between PD subtypes using our multidomain clinical criteria for subtyping, highlighting the role of multi-proteinopathies in people with PD with a faster disease progression.^4^

We found that participants with mild motor-predominant subtype showed further interval atrophy in the basal ganglia, likely indicating ongoing dopaminergic loss in the early stage, whereas in the diffuse-malignant subtype, the majority of dopaminergic cells are already damaged even in the early years as previously shown by more severe changes in baseline DAT scans in this PD subtype,^3^ suggesting a saturation effect. Nevertheless, the choice and sensitivity of the imaging modality are important determinants to depict between-subtype or between-stage differences in basal ganglia and/or substantia nigra integrity. Using 3-T MRI with 3-dimensional T1-weighted and neuromelanin-sensitive imaging, a previous study showed ongoing longitudinal change in volume and signal intensity of substantia nigra in both early- and advanced-PD patients after 2-3 years of follow-up.^17^

We then performed a similarity analysis to test the hypothesis whether these subtypes are pathologically distinct, or whether they represent the same pathological spectrum with the main difference being only in the speed of progression. Our findings demonstrated that the trajectory in overall pattern of brain atrophy toward that of the diffuse-malignant subtype was no greater in mild motor-predominant and intermediate subtypes after 3.5-years of follow-up than in healthy controls. Interestingly, only subjects with the diffuse-malignant subtype showed a significantly increasing within-group convergence over time. Taken together, these results support the presence of a distinct neuropathological pattern of progression, at least in a subgroup of people with more aggressive PD.

### Strengths and Limitations

Our study has several strengths that contribute to its robustness and clinical relevance. First, our analysis features one of the longest follow-up periods for comparing clinical trajectories between PD subtypes with >8 years of annual assessment, allowing for a greater understanding of disease progression within each subtype over time. Additionally, this is one the few studies to investigate longitudinal changes in brain atrophy using MRI, providing valuable insights into the structural alterations occurring in different PD subtypes. Methodologically, our study employs statistical models which enable us to adjust for confounding factors such as the aging effect and baseline clinical differences among participants with various PD subtypes. Moreover, our findings provide further evidence to support the biological and prognostic relevance our multi-domain clinical subtyping method for PD, offering a practical framework that clinicians can utilize in office settings to assign individuals to distinct subtypes with prognostic implications. This proof of concept enhances the clinical utility of our findings, paving the way for personalized management strategies tailored to the specific needs of PD patients. We acknowledge some study limitations. One of the main constraints of the study stems from the small sample size of participants with follow-up MRI, especially among those with the diffuse-malignant subtype, and the relatively short MRI follow-up time. While our research question was hypothesis-driven, and the results are statistically significant, future studies with a larger sample size will increase the confidence and generalizability of our findings. Furthermore, external validation in other PD cohorts is needed to further confirm the generalizability and robustness of our results.

## CONCLUSION

Our study provides further evidence that multi-domain subtyping, based on initial motor and three key non-motor features of PD (i.e., RBD, autonomic disturbance, and early cognitive deficit), is a valid method to separate subgroups of PD. We now see evidence for more aggressive long-term disease progression due to a distinct pattern of pathology. We have demonstrated an accelerated atrophy pattern within several brain regions in diffuse-malignant PD subtype compared to the ‘mild motor-predominant’ group, distinguishable even after a rather short follow-up period of 3.5 years. People with the diffuse-malignant subtype showed the highest within-group similarity in the overall pattern of brain MRI, while other subtypes failed to show a meaningful convergence towards the diffuse-malignant subtype over time, compared to that of normal aging. These findings suggest the presence of a more diffuse multidomain neurodegenerative process in a subgroup of people with PD, favoring the existence of bioanatomical PD subtypes with diverse underlying pathophysiology. Our study has demonstrated brain MRI evidence to explain part of the diversity in people with idiopathic PD fostering the concept of precision medicine.

## Data Availability

All data produced in the present study are available upon reasonable request to the authors

